# Taller height and risk of coronary heart disease and cancer: a within-sibship Mendelian randomization study

**DOI:** 10.1101/2021.07.16.21260639

**Authors:** Laurence J Howe, Ben Brumpton, Humaira Rasheed, Bjørn Olav Åsvold, Cristen J Willer, George Davey Smith, Neil M Davies

## Abstract

Taller people have lower risk of coronary heart disease but higher risk of many cancers. Mendelian randomization studies in unrelated individuals have suggested that these relationships are potentially causal. However, Mendelian randomization estimates from samples of unrelated individuals are sensitive to demography (population stratification, assortative mating) and familial (indirect genetic) effects. Height could influence disease risk via anatomic and physiological effects of height (e.g., number of cells or the bore of arteries) or previous results may have been confounded by early-life environmental factors (e.g., parental socioeconomic position and nutrition).

In this study, we performed within-sibship Mendelian randomization analyses using 77,757 siblings, a design robust against demography and indirect genetic effects of parents. Within-sibship Mendelian randomization estimated that one SD taller height lowers odds of coronary heart disease by 14% (95% CI: 3% to 23%) but increases odds of cancer by 18% (95% CI: 3% to 34%). There was some evidence that taller height reduces systolic blood pressure and LDL cholesterol, which may mediate some of the protective effect of taller height on coronary heart disease risk.

For the first time, we have demonstrated that purported effects of height on adulthood disease risk are unlikely to be explained by demographic or familial factors, and so likely reflect an individual-level causal effect. Disentangling the mechanisms via which height affects disease risk may improve understanding of the aetiologies of atherosclerosis and carcinogenesis.

## Introduction

Height is a classical complex trait influenced by genetic and early-life environmental factors. Despite the non-modifiable nature of adult height, evaluating the effects of height on non-communicable disease risk can give insights into the aetiology of adulthood diseases ^1 2^. Two major groupings of disease, cardiovascular disease and cancer, have divergent associations with height ^1-3^. Taller people are less likely to develop cardiovascular disease, including coronary heart disease ^1 4-8^ and stroke ^9^, but more likely to be diagnosed with cancer ^1 10-16^. The mechanisms via which height influences disease risks are unclear. The association between height and cardiovascular disease may be mediated via favourable lipid profiles ^1 4^, lower systolic blood pressure (SBP) ^1 17^, lung function ^8 18^, lower heart rate ^19^ and coronary artery vessel dimension ^20^. The increased cancer incidence amongst taller individuals could relate to early life exposure to hormones such as insulin-like growth factor 1 (IGF-1) ^21 22^ or the increased number of cells in taller individuals ^3 10 23^. However, although overall cancer risk is higher amongst taller individuals ^10 14 15^, there is some evidence for heterogeneity across cancer subtypes with null or inverse associations observed between height and risk of stomach, oropharyngeal and oesophageal cancers ^10 14-16^.

Height is highly heritable but the average height across European populations has increased over the last hundred years ^24^, illustrating effects of early-life environmental factors such as nutrition and childhood infections. The associations of height with adulthood diseases and relevant biomarkers could reflect biomechanical effects relating to increased stature (e.g. number of cells or larger arteries ^20^) or could reflect confounding by early-life environmental factors that influence both height and later-life health such as parental socio-economic position. For example, wealthier parents may provide their offspring with better nutrition, leading to increased adult height, and a better education, potentially leading to improved health in adulthood ^16^. Thus, it is unclear whether height has a causal effect on risk of cardiovascular disease and cancer or if a confounding factor influences both height and disease risk.

Mendelian randomization ^25^ analyses, using genetic variants associated with height as a proxy for observed height, have been used to strengthen the evidence for causal effects of height on adulthood diseases ^4 5 11-13^. The underlying premise being that genetic variants associated with height, unlike height itself, are unlikely to be associated with potential confounders such as childhood nutrition. However, there is growing evidence that estimates from genetic epidemiological studies using unrelated individuals may capture effects relating to demography (population stratification, assortative mating) and familial effects (e.g. indirect genetic effects of relatives where parental genotype influences offspring phenotypes) ^26-34^. Indeed, recent articles have illustrated the potential for genetic analyses of height to be affected by these biases ^27 28^ including a Mendelian randomization study of height on education ^31^. One approach to overcome these potential biases is to use data from siblings ^31 35^, and exploit the shared early-life environment of siblings and the random segregation of alleles during meiosis ^25^. Indeed, true Mendelian randomization was initially proposed as existing within a parent-offspring design ^25 36^ (**Figure 1**).

**Figure 1.**
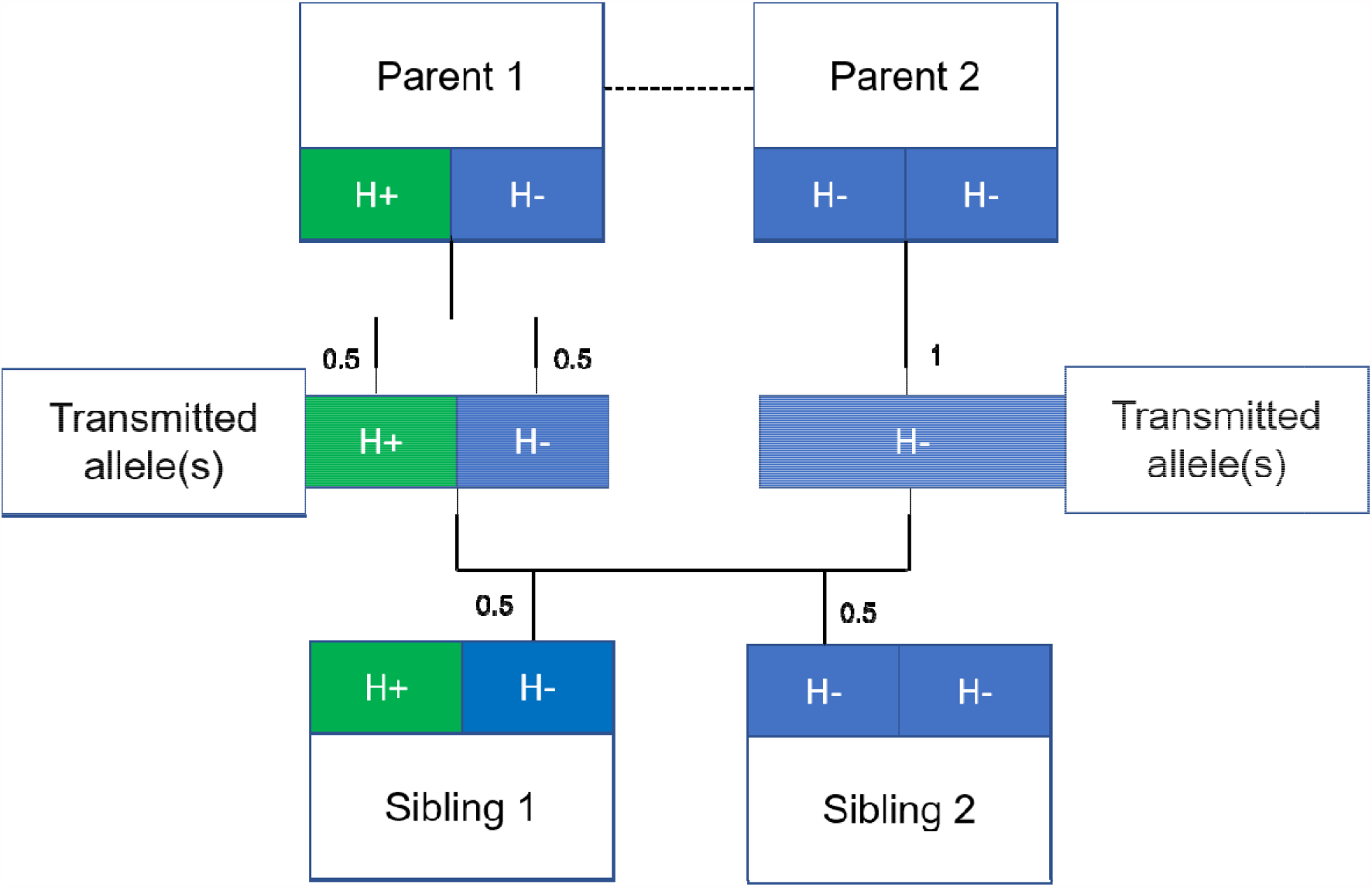
Mendelian randomization within families

Figure 1 illustrates the random allocation of alleles within a parent-offspring quad (two parents and two offspring), initially observed by Mendel. Consider a height influencing genetic variant H where on average individuals with the H+ allele are taller than individuals with the H-allele. From Mendel’s law of segregation, parent 1 who is heterozygous at this allele has equal chance of transmitting either a H+ or H-allele to offspring. Parent 2, homozygous at this allele, will always transmit a copy of the H-allele. It follows that 50% of this pair’s offspring will be heterozygous (as Parent 1) and 50% will be homozygous for the H-allele (as Parent 2). On average, the heterozygous offspring will be taller than the homozygous H-offspring, with this difference a consequence of random segregation of gametes.

Here we used data from 40,068 siblings from UK Biobank ^37^ and 37,689 siblings from the Norwegian HUNT study ^38^ to estimate effects of adulthood height on coronary heart disease (CHD), cancer risk and relevant biomarkers. We report estimates of the effects of height on coronary heart disease and cancer from both phenotypic models and Mendelian randomization, with and without accounting for family structure.

## Methods

### UK Biobank

#### Overview

UK Biobank is a large-scale prospective cohort study, described in detail previously ^37 39^. In brief, 503,325 individuals aged between 38-73 years were recruited between 2006 and 2010 from across the United Kingdom. For the purposes of this study, we used a subsample of 40,068 siblings from 19,523 families ^31^. Full-siblings were derived using UK Biobank provided estimates of pairwise identical by state (IBS) kinships (>0.5-21^*^IBS0, <0.7) and IBS0 (>0.001, <0.008), the proportion of unshared loci ^40^.

#### Phenotype data

At baseline, study participants attended an assessment centre where they completed a touch screen questionnaire, were interviewed, and had various measurements and samples taken. Height (field ID: 12144-0.0) and sitting height (field ID: 20015-0.0) were measured using a Seca 202 device at the assessment centre. Seated height is equivalent to trunk length, leg length was defined as height minus seated height and leg to trunk ratio was calculated by taking the ratio of leg and trunk length. SBP was measured using an automated reading from an Omron Digital blood pressure monitor (field ID: 4080-0.0). Biomarkers of interest including direct LDL cholesterol (LDL-C), HDL cholesterol (HDL-C), triglycerides (TG), glucose and insulin-like growth factor 1 (IGF-1) were measured using blood samples and the Beckman Coulter AU5800 or the DiaSorin LIASON XL (IGF-1) analysers.

International Classification of Disease (10^th^ edition) (ICD10) codes and Office of Population Censuses and Surveys Classifications of Interventions and Procedures (OPCS) codes were used to identify CHD and cancer (all subtypes and a stratified analysis) cases using several data sources; a) secondary care data from Hospital Episode Statistics (HES); b) death register data; and c) cancer registry data.

Relevant codes are contained in **Supplementary Table 5**.

#### Genotyping

UK Biobank study participants (N= 488,377) were genotyped using the UK BiLEVE (N= 49,950) and the closely related UK Biobank Axiom^™^ Arrays (N= 438,427). Directly genotyped variants were pre-phased using SHAPEIT3 ^41^ and imputed using Impute4 and the UK10K ^42^, Haplotype Reference Consortium ^43^ and 1000 Genomes Phase 3 ^44^ reference panels. More detail is contained in a previous publication ^37^.

### HUNT

#### Overview

The Trøndelag Health Study (HUNT) is a series of general health surveys of the adult population of the demographically stable Nord-Trøndelag region, Norway, as detailed in a previous study ^45^. The entire adult population of this region (∼90,000 adults in 1995) is invited to attend a health survey (includes comprehensive questionnaires, an interview, clinical examination, and detailed phenotypic measurements) every 10 years. To date four health surveys have been conducted, HUNT1 (1984–1986), HUNT2 (1995–1997), HUNT3 (2006–2008) and HUNT4 (2017-2019) and all surveys have high participation rate ^46^. This current study includes 38,723 siblings from 15,179 families who participated in the HUNT2 and HUNT3 surveys. Siblings were identified using KING software ^47^, with pairs defined as follows; kinship coefficient between 0.177 and 0.355, the proportion of the genomes that share two alleles IBD > 0.08, and the proportion of the genome that share zero alleles IBD > 0.04.

#### Phenotype data

Height was measured to the nearest 1.0 cm using standardized instruments with participants wearing light clothes without shoes. SBP was measured using automated oscillometry (Critikon Dinamap 845XT and XL9301, acquired by GE Medical Systems Information Technologies in 2000) on the right arm in a relaxed sitting position ^45 46^. SBP was measured twice with a one-minute interval between measurement with the mean of both measurements used in this study.

All HUNT participants provided non-fasting blood samples when attending the screening site. Total cholesterol, HDL-C and TG levels in HUNT2 were measured in serum samples using enzymatic colorimetric methods (Boehringer Mannheim, Mannheim, Germany). In HUNT3, participants’ total cholesterol was measured by enzymatic cholesterol esterase methodology; HDL-Cl was measured by accelerator selective detergent methodology; and TG were measured by glycerol phosphate oxidase methodology (Abbott, Clinical Chemistry, USA). LDL-C levels were calculated using the Friedewald formula ^48^ in both surveys. Participants in HUNT with TG levels ≥4.5mmol/L (n=1349) were excluded for LDL-C calculation, as the Friedewald formula is not valid at higher TG levels. For all these phenotypes, if the participant attended both HUNT2 and HUNT3 survey then values from HUNT2 were used for the analysis presented here.

The unique 11-digit identification number of every Norwegian citizen was used to link the HUNT participant records with the hospital registry, which included the three hospitals in the area (up to March 2019). We used ICD-10 and ICD-9 (International Classification of Disease-9 and -10) codes 410-414 and I20-I25 to define coronary heart disease. Cancer status (yes/no) was self-reported in HUNT2, HUNT3 and HUNT4 questionnaires. Individuals with discordant responses across different questionnaires were excluded from analyses.

#### Genotyping

DNA samples were available from 71,860 HUNT samples from HUNT2 and HUNT3 and were genotyped^46^ using one of the three different Illumina HumanCoreExome arrays: HumanCoreExome12 v1.0 (n= 7570), HumanCoreExome12 v1.1 (n=4960) and University of Michigan HUNT Biobank v1.0 (n=58041; *HumanCoreExome*-*24* v1.0, with custom content). Quality control was performed separately for genotype data from different arrays. The call rate of genotyped samples was >99%. Imputation was performed on samples of recent European ancestry using Minimac3 (v2.0.1, http://genome.sph.umich.edu/wiki/Minimac3)^49^ from a merged reference panel constructed from i) the Haplotype Reference Consortium panel (release version 1.1)^43^ and ii) a local reference panel based on 2,202 whole-genome sequenced HUNT participants^50^. Subjects included in the study were of European ancestry and had passed the quality control.

### Statistical analysis

#### Population and within-sibship models

The population model is a conventional regression model where the outcome is regressed (linear or logistic) against the exposure (height or height polygenic score (PGS)) with the option to include covariates.

The within-sibship model is an extension to the population model which includes a family mean term, the average exposure value across each family (height or height PGS), with each individual exposure value centred about the family mean exposure. To account for relatedness between siblings, standard errors are clustered by family in both models. More information on these models is contained in previous publications ^31 51^.

#### Phenotypic and Mendelian randomization analyses

In phenotypic analyses, we used regression models (within-sibship and population) to estimate the association between measured height and all outcomes (CHD, cancer, SBP, LDL-C, HDL-C, TG, glucose and IGF-1) using linear models for continuous outcomes and logistic models for binary disease outcomes. In both cohorts, we used a standardised measure of height after adjusting for age and sex and also standardised continuous outcomes after adjusting for age and sex.

In Mendelian randomization analyses, we fit regression models as above but used an age/sex standardised height PGS instead of measured height. The height PGS was constructed in PLINK ^52^ using 372 independent (LD clumping: 250 kb, r2 < 0.01) genetic variants from a previous height Genome-wide association study (GWAS) ^53^ which did not include UK Biobank or HUNT. Again, we standardised and adjusted for age/sex for continuous outcomes. To estimate the effect of the PGS on height, we fit a model regressing measured standardised height against the height PGS. We then generated scaled Mendelian randomization estimates by taking the Wald ratio of the PGS-outcome associations and the PGS-height associations. All statistical analyses were conducted using R (v 3.5.1).

There are three core instrumental variable assumptions for Mendelian randomization analyses. First, the genetic variants should be robustly associated with the exposure (relevance). Second, there should be no unmeasured confounders of the genetic variant-outcome association (independence). Third, the genetic variants should only influence the outcome via their effect on the exposure (the exclusion restriction) ^54-56^.

#### UK Biobank and HUNT meta-analyses

We performed phenotypic and Mendelian randomization analyses (using population and within-sibship models) in both UK Biobank and HUNT. For phenotypes measured in both studies (CHD, cancer, LDL-C, HDL-C, TG), we combined estimates across both studies using a fixed-effects model in the metafor R package for meta-analysis. We tested for heterogeneity between UK Biobank/HUNT estimates using the difference of two means test statistic ^57^.

#### Outcomes

Using the previously described models and meta-analysis procedure, we estimated the effects of height on CHD, cancer, LDL-C, HDL-C, TG, glucose and IGF-1. As a sensitivity analysis, we used phenotypic models to evaluate associations between dimensions of height (leg length, trunk length and leg to trunk ratio) with CHD and cancer in UK Biobank. A further sensitivity analysis involved repeating cancer analyses in UK Biobank with a subset of cancers not phenotypically associated with height (described above).

## Results

### Adulthood height and risk of coronary heart disease and cancer

We found consistent evidence across population and within-sibship models, using both measured height and a height PGS, that taller adulthood height reduced CHD risk and increased risk of cancer (**Supplementary Tables 1/2**).

Within-sibship Mendelian randomization estimated that 1 SD taller height (approximately 6.8 cm for men and 6.2cm for women) reduced odds of CHD by 14% (95% C.I. 3% 23%) but increased the odds of cancer by 18% (95% C.I. 3%, 34%). These estimates were consistent across analyses using measured height as well as with population Mendelian randomization estimates. For example, population Mendelian randomization analyses estimated that 1 SD taller height reduced odds of CHD by 10% (95% C.I. 4%, 16%) and increased odds of cancer by 9% (95% C.I. 2%, 16%) (**Table 1**/ **Figure 2**).

**Table 1.**
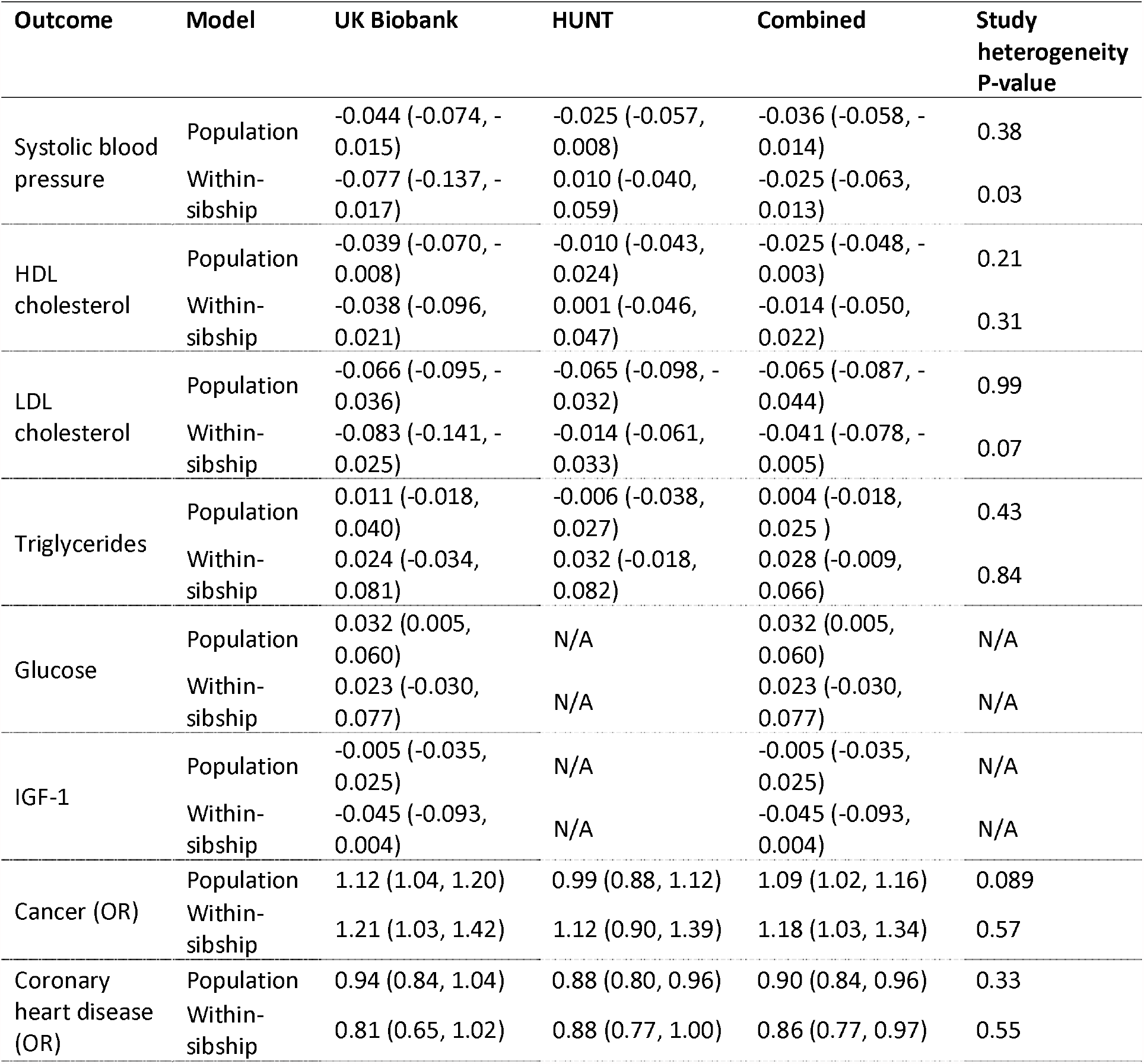
Mendelian randomization results: Change in outcome (SD units), per 1 S.D. increase in height.

**Figure 2.**
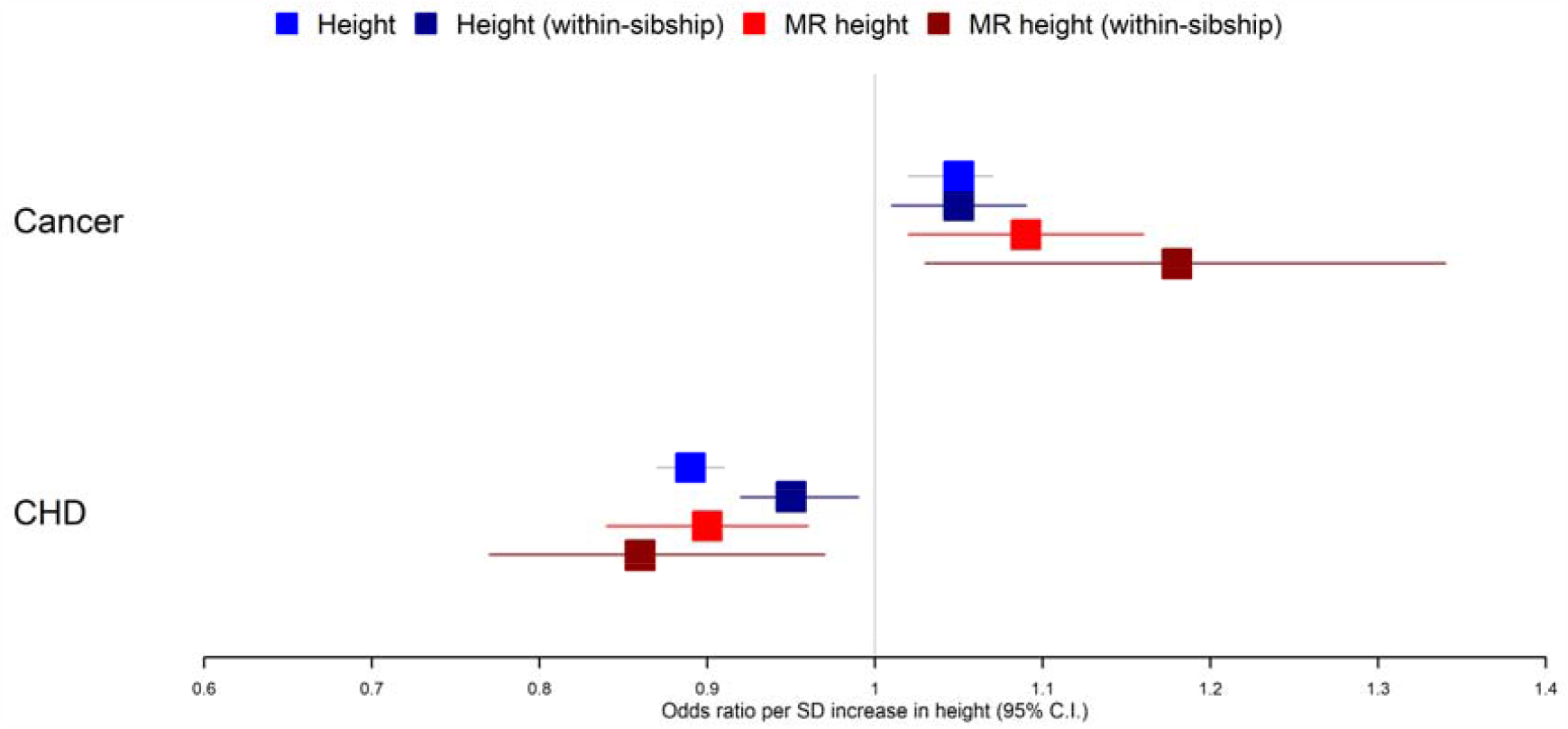
Taller height and risk of coronary heart disease and cancer

Figure 2 displays meta-analysis results from 4 different models used to evaluate the effect of height on CHD and cancer risk. First, a phenotypic population model with measured height as the exposure and age and sex included as covariates. Second, a within-sibship phenotypic model with the family-mean height included as an additional covariate to account for family structure. Third, a population Mendelian randomization model with height PGS as the exposure exploiting advantageous properties of genetic instruments. Fourth, a within-sibship Mendelian randomization model with the family mean PGS included as a covariate to control for parental genotypes. Across all 4 models, we found consistent evidence that taller height reduces odds of CHD and increases odds of cancer.

We then evaluated associations between dimensions of height (trunk length, leg length and leg to trunk ratio) and risk of CHD/cancer in UK Biobank. We found little evidence of heterogeneity between estimates although stronger conclusions are limited by statistical power (**Supplementary Table 3**). We also ran a sensitivity analysis in UK Biobank, rerunning height-cancer analyses including only cases with one of 7 cancer subtypes (lung, oropharyngeal, stomach, oesophageal, pancreatic, bladder and multiple myeloma) for which a previous study found little evidence they associated with height ^10^. These subtypes generally show very strong social patterning which could explain the attenuated associations with height which is also often socially patterned. As expected, the association of measured height with this subset of cancers (population OR 0.99; 95% C.I. 0.92, 1.06; within-sibship OR 1.01; 95% C.I. 0.88, 1.15) was less strong than the association between height and the all-cancer outcome (population OR 1.05; 95% C.I. 1.02, 1.07; within-sibship OR 1.05; 95% C.I. 1.01, 1.09). Mendelian randomization estimates were imprecise because of the modest number of cases for these cancers (**Supplementary Table 4**).

### Adulthood height and biomarkers

Using measured biomarkers, both population and within-sibship models found evidence for association between taller height and lower SBP, lower circulating LDL-C and higher circulating IGF-1 levels. There was some evidence for heterogeneity in phenotypic associations between height and biomarkers in UK Biobank and HUNT, such as for SBP, which was more strongly associated with height in UK Biobank (**Supplementary Table 1**).

Population Mendelian randomization results suggested that taller height reduced SBP (per 1 SD taller height, 0.036 SD decrease; 95% C.I. 0.014, 0.058), LDL-C (per 1 SD taller height, 0.065 SD decrease; 95% C.I. 0.044, 0.087), HDL-C (per 1 SD taller height, 0.025 SD decrease; 95% C.I. 0.003, 0.048) but increased glucose (per 1 SD taller height, 0.032 SD increase; 95% C.I. 0.005, 0.060). In contrast, we found little evidence that taller height affected TG or IGF-1 levels. Within-sibship Mendelian randomization estimates were consistent with population estimates; SBP (per 1 SD taller height, 0.025 SD decrease; 95% C.I. -0.013, 0.063), LDL-C (per 1 SD taller height, 0.041 SD decrease; 95% C.I. 0.005, 0.078), HDL-C (per 1 SD taller height, 0.014 SD decrease; 95% C.I. -0.022, 0.050) and glucose (per 1 SD taller height, 0.023 SD increase; 95% C.I. -0.030, 0.077) (**Figure 3 / Table 1**).

**Figure 3.**
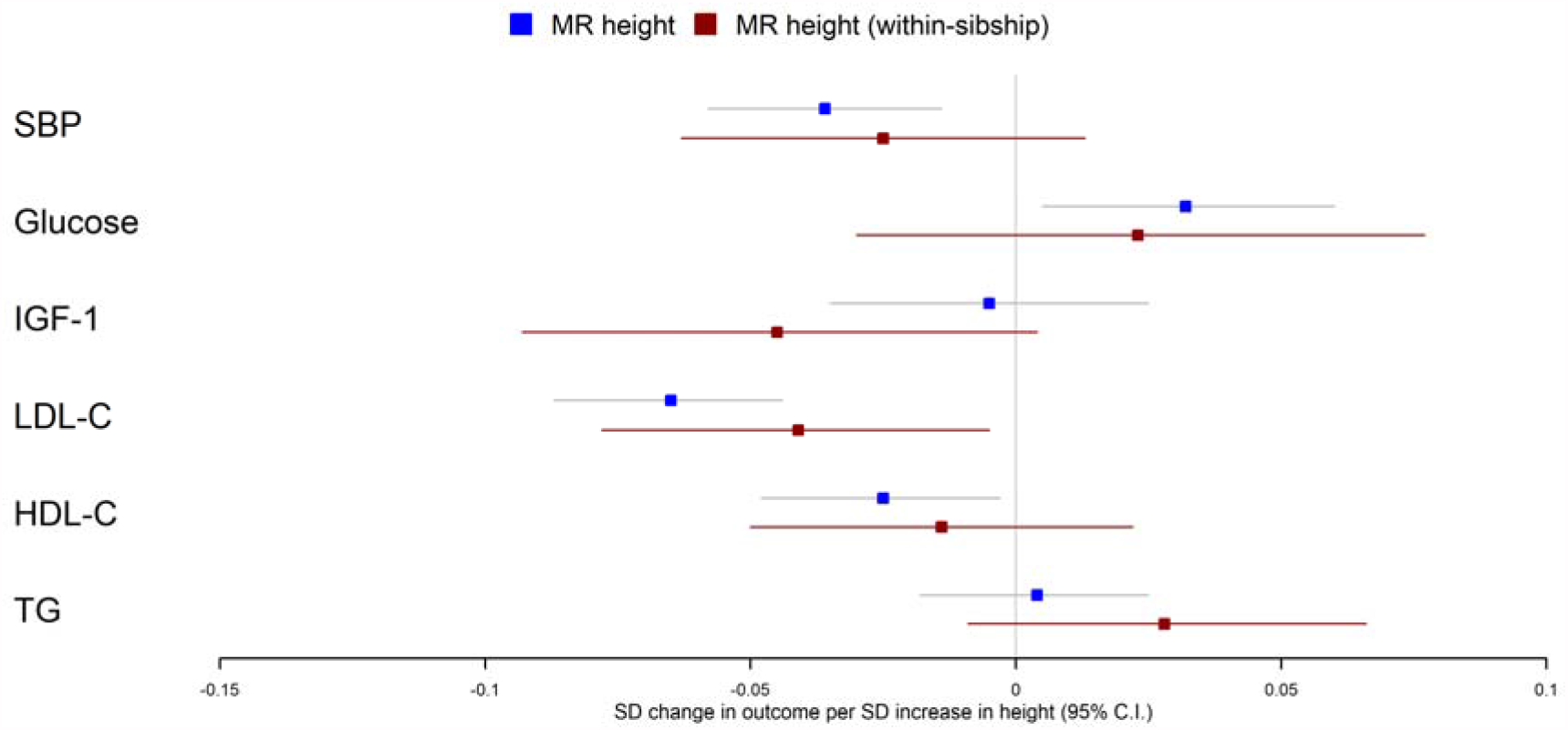
Mendelian randomization estimates of effects of taller height on biomarkers

There was some putative evidence for heterogeneity in Mendelian randomization effect estimates between UK Biobank and HUNT. For example, within-sibship Mendelian randomization estimate suggested effects of height on SBP in UK Biobank (0.077 SD decrease; 95% C.I. 0.017, 0.137) but the effect estimate was in the opposite direction in HUNT (0.010 SD increase; 95% C.I. -0.040, 0.059; heterogeneity P = 0.03) (**Table 1**).

Figure 3 shows meta-analysis results from population and within-sibship Mendelian randomization analyses estimating the effect of taller height on biomarkers across UK Biobank and HUNT. The estimates were broadly similar between the two models suggesting modest effects of demography and indirect genetic effects.

## Discussion

In this study, we used sibling data from two large biobanks to estimate the effects of height on CHD, cancer and relevant biomarkers. We found consistent evidence across all models, including within-sibship Mendelian randomization, that taller height is protective against CHD but increases risk of cancers. We found less consistent evidence for effects of height on biomarkers; population and within-sibship phenotypic models as well as population Mendelian randomization models suggested modest effects of taller height on SBP, LDL-C and HDL-C. However, the confidence intervals for within-family Mendelian randomization of height and biomarkers were too wide to draw strong conclusions.

Our findings are largely consistent with previous studies ^1 4-6 8 10-13 58^, that used non-sibling designs, and with the hypothesis that height affects CHD and cancer risk. However, previous studies were potentially susceptible to bias relating to geographic and socioeconomic variation in height and height genetic variants ^26 28 32^. Indeed, a recent within-sibship Mendelian randomization study found that previously reported effects of height and body mass index on educational attainment greatly attenuated when using siblings ^31^. Here, we provided robust evidence for individual-level effects of height by demonstrating that previous evidence for effects of height on adulthood disease risk is unlikely to have been confounded by demography or indirect genetic effects. Major strengths of our work are the use of within-sibship Mendelian randomization ^35^, and the triangulation ^59^ of evidence from across phenotypic, genetic and within-sibship models. A major limitation of our analyses is that because of limited sibling data and the statistical inefficiency of within-family models, we have limited statistical power to investigate effects of height on disease subtypes and to further explore mechanisms using multivariable Mendelian randomization ^60^.

Adulthood height is non-modifiable and the interpretation of causality is nuanced because it is unclear whether biological effects relate to stature itself, increased childhood growth or to factors highly correlated with height such as lung function ^8 18^ and artery length ^61^. Previous studies ^15 17 18 62^ have explored the possibility that associations may relate to dimensions of height, with evidence that blood pressure is associated with trunk but not leg length ^62^. Here, we found that the effects of height on disease risk due to leg or trunk length were similar. We found consistent effects of increased height across aetiologically heterogeneous cancer subtypes, which implies that the mechanism could relate to the larger number of cells in taller individuals or a generalized growth phenotype. Our Mendelian randomization estimates for effects of height on a subset of cancers not strongly phenotypically associated with height ^10^ were consistent with the combined cancer estimates, although we had limited power in this dataset because of the modest prevalence of the cancer subtypes.

The estimated effects of height on disease risk were relatively consistent between the Norwegian HUNT and UK Biobank studies. Contrastingly, the heterogeneity between UK Biobank and HUNT for analyses involving SBP and LDL-C suggests that some effects of height could be population specific. Alternatively, heterogeneity could relate to the variance in associations between adulthood height and early-life environmental confounders across countries ^16^. Additional explanations could relate to differences in biomarker measurement between studies (e.g. measuring LDL-C directly or using the Friedewald formula, differences in fasting level before samples were taken) or selection bias ^63^.

To conclude, using within-sibship Mendelian randomization, we showed that height has individual-level effects on risk of CHD and cancers as well as several biomarkers. Larger family datasets and additional analyses including two-step ^64^ and multivariable Mendelian randomization ^60^ could be used to investigate potential mediators of these relationships.

## Data Availability

UK Biobank individual level participant data are available via enquiry to. If interested in accessing HUNT data, you can find more information at https://www.ntnu.edu/hunt/data. Example scripts for population and within-sibship models are available on GitHub https://github.com/LaurenceHowe/SiblingGWAS.

## Acknowledgements

This research has been conducted using the UK Biobank Resource under Application Number 15825. UK Biobank has ethical approval from the North West Multi-centre Research Ethics Committee (MREC). Quality Control filtering of the UK Biobank data was conducted by R.Mitchell, G.Hemani, T.Dudding, L.Paternoster as described in the published protocol(doi:10.5523/bris.3074krb6t2frj29yh2b03x3wxj). The University of Bristol support the MRC Integrative Epidemiology Unit [MC_UU_00011/1]. NMD is supported by a Norwegian Research Council Grant number 295989.

The Trøndelag Health Study (The HUNT Study) is a collaboration between HUNT Research Centre (Faculty of Medicine and Health Sciences, NTNU, Norwegian University of Science and Technology), Trøndelag County Council, Central Norway Regional Health Authority, and the Norwegian Institute of Public Health. The funders had no role in study design, data collection and analysis, decision to publish, or preparation of the manuscript. This publication is the work of the authors, who serve as the guarantors for the contents of this paper.

## Contributions

LJH formulated the project, drafted the manuscript, and performed statistical analyses under the supervision of NMD and GDS.

HR and BB prepared datasets and performed QC in the HUNT study.

All authors contributed to interpretation of results and drafting of the final manuscript.

