## Supplementary Tables for "Taller height and risk of coronary heart disease and cancer: a within-sibship Mendelian randomization study"

**Supplementary Table 1** Phenotypic results: Change in outcome (SD units), per 1 S.D. increase in height.

| **Outcome** | **Model** | **UK Biobank** | **HUNT** | **Combined** | **Study heterogeneity**  **P-value** |
| --- | --- | --- | --- | --- | --- |
| Systolic blood pressure | Population | -0.066 (-0.076, -0.056) | -0.040 (-0.051, -0.029) | -0.054 (-0.061, -0.046) | < 0.001 |
|  | Within-sibship | -0.042 (-0.061, -0.024) | -0.006 (-0.022, 0.010) | -0.021 (-0.033, -0.009) | 0.004 |
| HDL cholesterol | Population | 0.013 (0.002, 0.024) | -0.008 (-0.019, 0.003) | 0.003 (-0.005, 0.011) | 0.010 |
|  | Within-sibship | -0.030 (-0.048, -0.012) | -0.031 (-0.046, -0.015) | -0.030 (-0.042, -0.019) | 0.95 |
| LDL cholesterol | Population | -0.018 (-0.029, -0.008) | -0.033 (-0.044, -0.022) | -0.025 (-0.033, -0.018) | 0.059 |
|  | Within-sibship | -0.022 (-0.040, -0.003) | -0.030 (-0.046, -0.015) | -0.027 (-0.039, -0.015) | 0.47 |
| Triglycerides | Population | -0.044 (-0.054, -0.034) | -0.014 (-0.026 -0.003) | -0.030 (-0.038, -0.023) | < 0.001 |
|  | Within-sibship | -0.010 (-0.028, 0.008) | 0.005 (-0.012, 0.022) | -0.002 (-0.015, 0.010) | 0.24 |
| Glucose | Population | -0.018 (-0.029, -0.007) | N/A | -0.018 (-0.029, -0.007) | N/A |
|  | Within-sibship | -0.008 (-0.029, 0.011) | N/A | -0.008 (-0.029, 0.011) | N/A |
| IGF-1 | Population | 0.070 (0.060, 0.081) | N/A | 0.070 (0.060, 0.081) | N/A |
|  | Within-sibship | 0.050 (0.032, 0.068) | N/A | 0.050 (0.032, 0.068) | N/A |
| Cancer (OR) | Population | 1.10 (1.07, 1.13) | 1.00 (0.96, 1.04) | 1.05 (1.02, 1.07) | 0.006 |
|  | Within-sibship | 1.09 (1.04 1.15)) | 1.04 (0.96, 1.11) | 1.05 (1.01, 1.09) | 0.71 |
| Coronary heart disease (OR) | Population | 0.83 (0.80, 0.86) | 0.92 (0.89, 0.94) | 0.89 (0.87, 0.91) | < 0.001 |
|  | Within-sibship | 0.86 (0.80, 0.92) | 0.98 (0.94, 1.03) | 0.95 (0.92, 0.99) | 0.001 |

**Supplementary Table 2** Change in outcome (SD units), per 1 S.D. increase in height PGS.

| **Outcome** | **UK Biobank** | |  | **HUNT** | |
| --- | --- | --- | --- | --- | --- |
|  | **Population** | **Within-sibship** |  | **Population** | **Within-sibship** |
| Height | 0.361 (0.350, 0.371) | 0.311 (0.298, 0.323) |  | 0.335 (0.324, 0.346) | 0.334 (0.321, 0.346) |
| Systolic blood pressure | -0.016 (-0.027, -0.005) | -0.024 (-0.042, -0.005) |  | -0.008 (-0.019, 0.003) | 0.003 (-0.013, 0.020) |
| HDL cholesterol | -0.014 (-0.025, -0.003) | -0.012 (-0.030, 0.006) |  | -0.003 (-0.14, 0.008) | 0.000 (-0.015, 0.016) |
| LDL cholesterol | -0.024 (-0.034, -0.013) | -0.026 (-0.044, -0.008) |  | -0.022 (-0.033, -0.011) | -0.005 (-0.020, 0.011) |
| Triglycerides | 0.004 (-0.006, 0.015) | 0.007 (-0.010, 0.025) |  | -0.002 (-0.13, 0.009) | 0.011 (-0.006, 0.027) |
| Glucose | 0.012 (0.002, 0.022) | 0.008 (-0.011, 0.028) |  | NA | NA |
| IGF-1 | -0.002 (-0.012, 0.009) | -0.016 (-0.034, 0.001) |  | NA | NA |
| Cancer (OR) | 1.04 (1.02 1.07) | 1.06 (1.01, 1.12) |  | 1.00 (0.96, 1.04) | 1.04 (0.96, 1.12) |
| Coronary heart disease (OR) | 0.98 (0.94, 1.01) | 0.94 (0.87, 1.01) |  | 0.96 (0.93, 0.98) | 0.96 (0.92, 1.00) |

**Supplementary Table 3** Associations of measured leg and trunk length with risk of cancer and coronary heart disease in UK Biobank

| **Outcome** | **Model** | **OR of outcome per SD increase (95% C.I.)** | | | |
| --- | --- | --- | --- | --- | --- |
|  |  | **Height* (REF)** | **Leg length** | **Trunk length** | **Leg/trunk ratio** |
| Cancer | Population | 1.10 (1.07, 1.13) | 1.09 (1.06, 1.12) | 1.07 (1.04, 1.10) | 1.04 (1.01, 1.06) |
|  | Within-sibship | 1.09 (1.04, 1.15) | 1.07 (1.02, 1.12) | 1.05 (1.01, 1.10) | 1.02 (0.97, 1.06) |
| Coronary heart disease | Population | 0.83 (0.80, 0.86) | 0.84 (0.81, 0.88) | 0.89 (0.86, 0.92) | 0.93 (0.90, 0.97) |
|  | Within-sibship | 0.86 (0.80, 0.92) | 0.89 (0.84, 0.96) | 0.92 (0.86, 0.98) | 0.97 (0.92, 1.04) |

* from UK Biobank only

**Supplementary Table 4** Height and cancer subtypes with limited evidence for associations with height in UK Biobank

| **Analysis** | **Model** | **OR of cancer per SD increase in height (95% C.I.)** | |
| --- | --- | --- | --- |
|  |  | **All cancers (REF: 9,146 cases)** | **Subset of cancers* in UK Biobank (909 cases)** |
| Phenotypic | Population | 1.05 (1.02, 1.07) | 0.99 (0.92, 1.06) |
|  | Within-sibship | 1.05 (1.01, 1.09) | 1.01 (0.88, 1.15) |
| Mendelian randomization | Population | 1.09 (1.02, 1.16) | 1.11 (0.93, 1.35) |
|  | Within-sibship | 1.18 (1.03, 1.34) | 1.23 (0.81, 1.86) |

* Cancers with limited evidence for associations with height (lung, oropharyngeal, stomach, oesophageal, pancreatic, bladder and multiple myeloma)

**Supplementary Table 5** Definitions of coronary heart disease and cancer in UK Biobank

| **Phenotype** | **Data source (codes)** |
| --- | --- |
| Coronary heart disease | Hospital Episode Statistics (ICD10: I21-I25, Z95)  Mortality registry (ICD10: I21-I25, Z95)  Hospital Procedures (OPCS: K40-K46, K471, K49, K50, K75) |
| Cancer (all cancers) | Hospital Episode Statistics (ICD10: Any C code)  Cancer registry (ICD10: Any C code)  Mortality registry (ICD10: Any C code) |
| Subset of cancers* | Hospital Episode Statistics (ICD10: C34, C0, C15-16, C25, C67, C90)  Cancer registry (ICD10: C34, C0, C15-16, C25, C67, C90)  Mortality registry (ICD10: C34, C0, C15-16, C25, C67, C90) |

* Cancers with limited evidence for associations with height (lung, oropharyngeal, stomach, oesophageal, pancreatic, bladder and multiple myeloma)
